# Effects of parity on the age at menopause and menopausal syndrome: a cross-sectional study in Northwest China

**DOI:** 10.1101/2020.04.18.20070706

**Authors:** Xiaoyan Sun, Weiguo Li, Rui Zhang, Lirong Wang, Xiping Shen, Yongbin Lu, Junxia An, Liyan Wang, Yiqing Wang, Xiaorong Luo, Haiying Zhu, Xuehong Zhang

## Abstract

**Objective:** This study evaluated the relationship between the number of births and the age at menopause and menopausal syndrome among Chinese women in Gansu.

**Methods:** A cross-sectional study was conducted by a local university from March to November in 2016. A total of 7236 women aged 40–55 years met study eligibility criteria. The modified KMI was used to assess the menopausal syndrome. Cox regression was applied to estimate HR and 95% CI of the relationship between parity and age at menopause. Logistic regression was performed to calculate OR and CI of the effects of birth times on the menopausal syndrome.

**Outcome measures:** The relationship between the parity and the age at menopause and menopausal syndrome in mid-life northwest Chinese women was analyzed.

**Results:** The mean age at menopause was 47.91 ± 3.31 years. The relationships between parity and age at menopause were not significant by applying Cox regression (P = 0.488). Women with nulliparity and more births (3 and ≥4) seemed to have higher risks of moderate and severe menopausal syndrome. In addition, the potential beneficial effects of 1 or 2 births on menopausal syndrome were still observed by applying the multivariable logistic regression analysis, particularly in urogenital symptoms.

**Conclusions:** No obvious relationship was found between parity and age at menopause among Chinese women in Gansu. Women with nulliparity and more births appeared to be at the increased risk of menopause syndrome compared with women with 1 and 2 births. The underlying mechanisms were not assessed and deserved further investigation.

**Highlight:** . The mean age at menopause was lower among Chinese women in Gansu than developed countries and regions.

. There is no obvious relationship between parity and age at menopause.

. Women with nulliparity and more births (3 and ≥4) appeared to be at the increased risk of menopause syndrome, particularly in urogenital symptoms.

## Introduction

Menopause is the hallmark of the loss of ovarian follicular activity, permanent cessation of menstruation, and loss of fertility, which is quite distressing and affects the personal, social, and work lives of women. The average age of menopause is about 50 years, whereas the mean lifetime is more than 80 years. Thus, women spend one third of their life in the menopause period. They may suffer from related symptoms or diseases due to an imbalance of endocrine system and deficiency of estrogen. Menopausal symptoms, especially severe menopausal symptoms, can harm women’s health and affect their daily life. Nevertheless, women with menopause have higher risks of metabolic syndrome[1], diabetes[2], and cardiovascular diseases[3].

The number of births affects the menopause. A previous study showed that the nulliparity lowered the age of menopause onset[4]. Meanwhile, a study about Shanghai women found that increased parity was associated with slightly later menopause and a longer reproductive span[5]. Some data suggested no relationship between parity and age at menopause occurrence[6]. However, most studies did not further analyze the relationship between them. Further, the information on Northwest China has not been reported yet.

Birth history influences women health and menopausal syndrome. Many studies reported that multiparity had an impact on the health of middle-aged women. Klingberg and colleagues showed that grand multiparous women (≥5 children) had increased risk of cardiovascular disease and myocardial infarction[7]. Another study also reported that the greater number of deliveries was a risk factor for health-related quality-of-life problems in South Korean women[8]. A recent study from Japan showed that the risk of type 2 diabetes tended to increase with the number of parity [9]. Other studies suggested that higher parity number was significantly associated with increased risk of cancer[10; 11].

Although the relationship between the number of births and menopause has been reported, the findings have not been consistent as described earlier. Therefore, a cross-sectional survey was performed to find out whether the number of births could affect the age at natural menopause and the frequency and intensity of menopausal syndrome among middle-aged women in Gansu Province of China. The study aimed to provide reproduction-related suggestions to women and protect their long-term health.

## Methods

### Study design and participants

This cross-sectional and observational study was conducted in China from 13 cities in Gansu Province, including 162 townships/towns in 54 counties/districts, using the multi-stage cluster random sampling method from March to November in 2016. The study inclusion criteria were as follows: (1) age 40–55 years; (2) residents living in the area for more than 1 year; (3) no use of hormonal drugs for at least 3 months; and (4) voluntary participation in the study and signed informed consent. The exclusion criteria were as follows: (1) menopause before the age of 40 years; (2) hearing disorder; (3) cognitive disorder; (4) unilateral or bilateral hysterectomy or ovariectomy at any time; (5) a history of radiotherapy or chemotherapy; (6)pregnant or breastfeeding; and (7) menopausal hormone therapy.

### Questionnaire and physician examination

The main aim of this study were to determine the impact of parity on age at menopause and menopausal syndrome. A questionnaire was designed, and it comprised (1) demographic data, including birth year, marital status, educational background attainment, employment status, and family monthly income; (2) details on menstrual and reproductive history, including age at menarche, age at first and last pregnancy, age at first and last childbearing, number of pregnancy and history of abortion, menopausal status, parity, breastfeeding, and menopausal age collected via personal interviews; and (3) modified Kupperman Index (KMI) scoring scale. Height and weight were measured and recorded. The modified KMI score to evaluate 13 menopausal symptoms included hot flashes and sweating, paresthesia, insomnia, irritability, depression, dizziness, fatigue, joint pain myalgia, sexual life abnormality, headache, palpitations, and urinary symptoms. They were divided into four grades (0–3 points) according to severity: 0, no symptoms; 1, mild; 2, moderate; and 3, severe. The weighted score was 4 for hot flashes and sweating; 2 points each for paresthesia, insomnia, irritability, sexual life abnormality, and urinary infection; and 1 point each for other symptoms. Total score = score of each symptom × weighted coefficient. The morbidity of menopausal syndrome was calculated using a KMI score cutoff of ≥7, which was the diagnostic criterion. The menopausal syndrome was considered mild with KMI scores of 7–15 points, moderate with KMI scores of 16–30, and severe with KMI scores of >30.

The KMI consisted of 13 items assessing menopausal symptoms, which were divided into the following 3 subscales: (1) somatic symptoms, including hot flashes, paresthesia, insomnia, vertigo, fatigue, muscle/joint pain, headache, palpitations, and formication; (2) psychological symptoms, including mood swings and melancholia; and (3) urogenital symptoms, including sexual and urinary problems.

### Criteria for premenopause, perimenopause, and postmenopause

According to the Stages of Reproductive Aging Workshop criteria) classification, the standard stages were as follows[12] :(1) premenopause characterized by minor changes in cycle length, particularly decreasing length of the menstrual cycle, and regular menstrual cycles with ≥12 menstruations during the last 12 months; (2) perimenopause characterized by a variable cycle length (persistent ≥7-day difference in the length of consecutive cycles), missed two or more cycles, and an episode of amenorrhea lasting more than 60 days during the last 12 months; and (3) postmenopause characterized by no menstrual bleeding during the last 12 months (at least 12 months of amenorrhea).

### Quality control

A self-administered questionnaire was designed by an epidemiologist (Rui Zhang) and a gynecological endocrinologist (Professor Xuehong Zhang). It comprised items spanning over eight pages and took about 30 min to complete. All investigators were uniformly trained on physical measurement methods, questionnaire explanation, and notes on completing the questionnaire. They passed the examination. All the interviewees completed the survey in a face-to-face manner. The questionnaire was recovered on the spot after completion and uniformly numbered in the order of site and recall questionnaire. The data were entered into the Epidata database by one investigator and one statistician. The consistency of data was checked. If they were found to be inconsistent, the original questionnaire was checked and modified.

### Estimation of sample size and statistical analysis

Data from previous studies suggested that 75%[13; 14] [15]of midlife women suffered from menopause syndrome. The parameters of incidence were set at 75%, accuracy at 0.05, and bilateral confidence intervals (CIs) at 95%. Further, the minimum sample size of each layer was considered as 128. The sample size was enlarged by 30% to reduce the sampling error, and the final number of women to be recruited was about 9000.

The statistical analysis was performed using SPSS 24.0 version. The continuous quantitative variables were expressed as mean ± standard deviation (x ± s). The qualitative variables were expressed as medians and percentages. The chi-square test and analysis of variance were used to compare categorical and continuous data, respectively. Non-normal distribution data were represented by median (25%, 75%), and nonparametric tests were used. Median and mean age at natural menopause were evaluated using nonparametric Kaplan–Meier cumulative survivorship estimates. Cox regression was applied to estimate the hazard ratio (HR) and 95% CI of the relationship between parity and age at menopause, and a logistic regression model was used to calculate the odds ratio (OR) and CI of the effects of birth times on the menopausal syndrome. A *P* value <0.05 indicated a statistically significant difference.

## Results

A total of 8500 women agreed to enroll in the study, and 8446 questionnaires were reclaimed. Further, 508 were ineffective questionnaires, including 379 cases of incomplete demographic information,67 cases of age <40 years, and 62 cases of similar surveys, and hence removed. In addition, 702 questionnaires were excluded for reasons of surgical menopause and incomplete or error information on menstruation and birth history. A total of 7236 women who completed the questionnaires were included in the study. The effective response rate was 85.67%.

### Characteristics of the study population

The baseline demographics of the 7236 participants are summarized in Table 1. Further, 60 were nulliparous women; 3500 had 2 births, and 327 had ≥4 births, accounting for 48.40% and 4.46%, respectively. The rates of women educated beyond high school with 3 and 4 births were significantly higher than the rates of those with 1 birth (92.65% and 97.86% vs 44%, *P* = 0.000). The proportions of low income and residence in rural areas were the highest in women with ≥4 births (53.82%, *P* = 0.000 and 66.06%, *P* = 0.000, respectively). The age at first delivery was significantly lower in women with ≥4 births than in those with 1 birth (21.39 ± 2.32 vs 25.54 ± 2.88, *P* = 0.000).

**Table 1.**
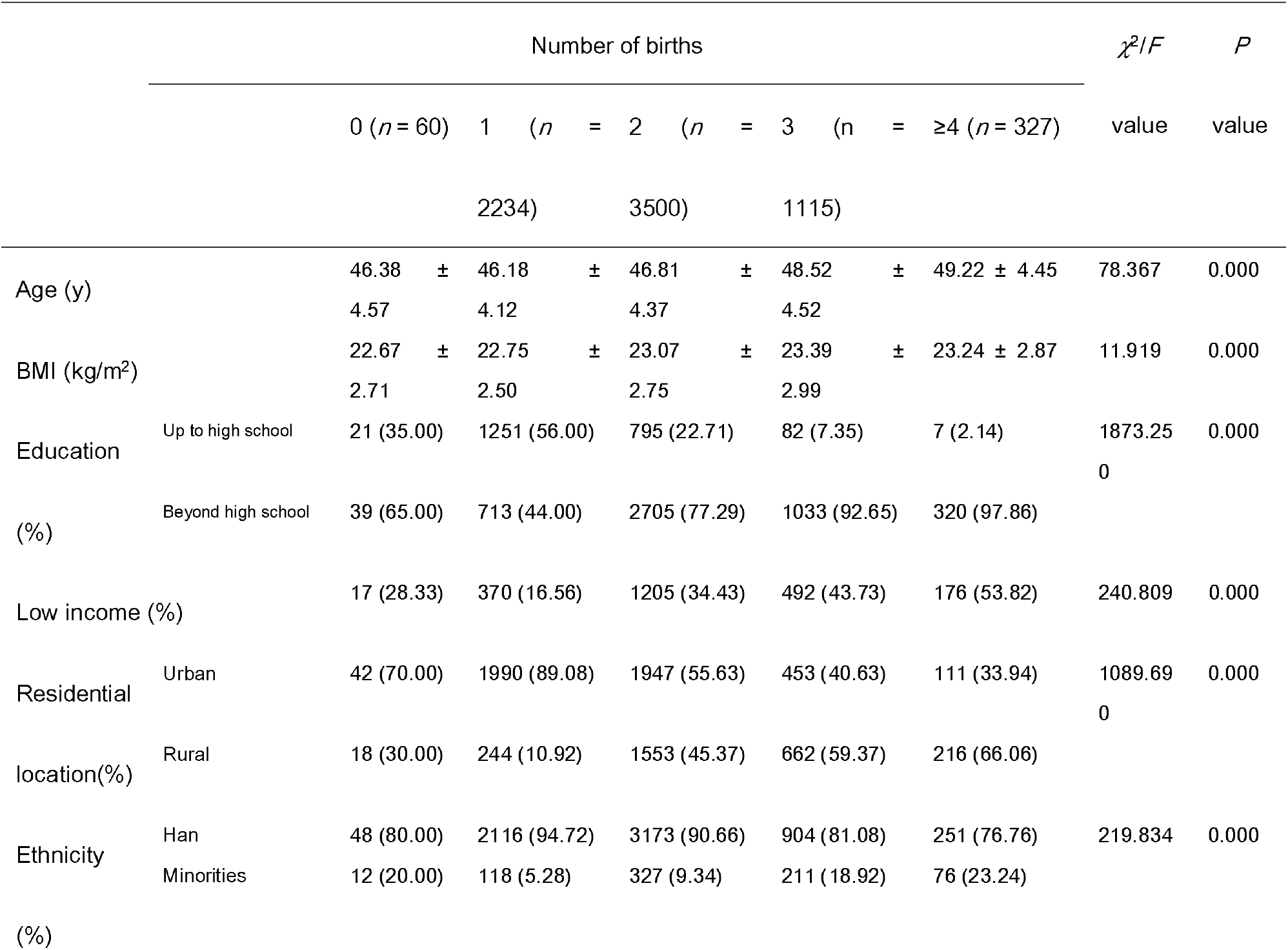
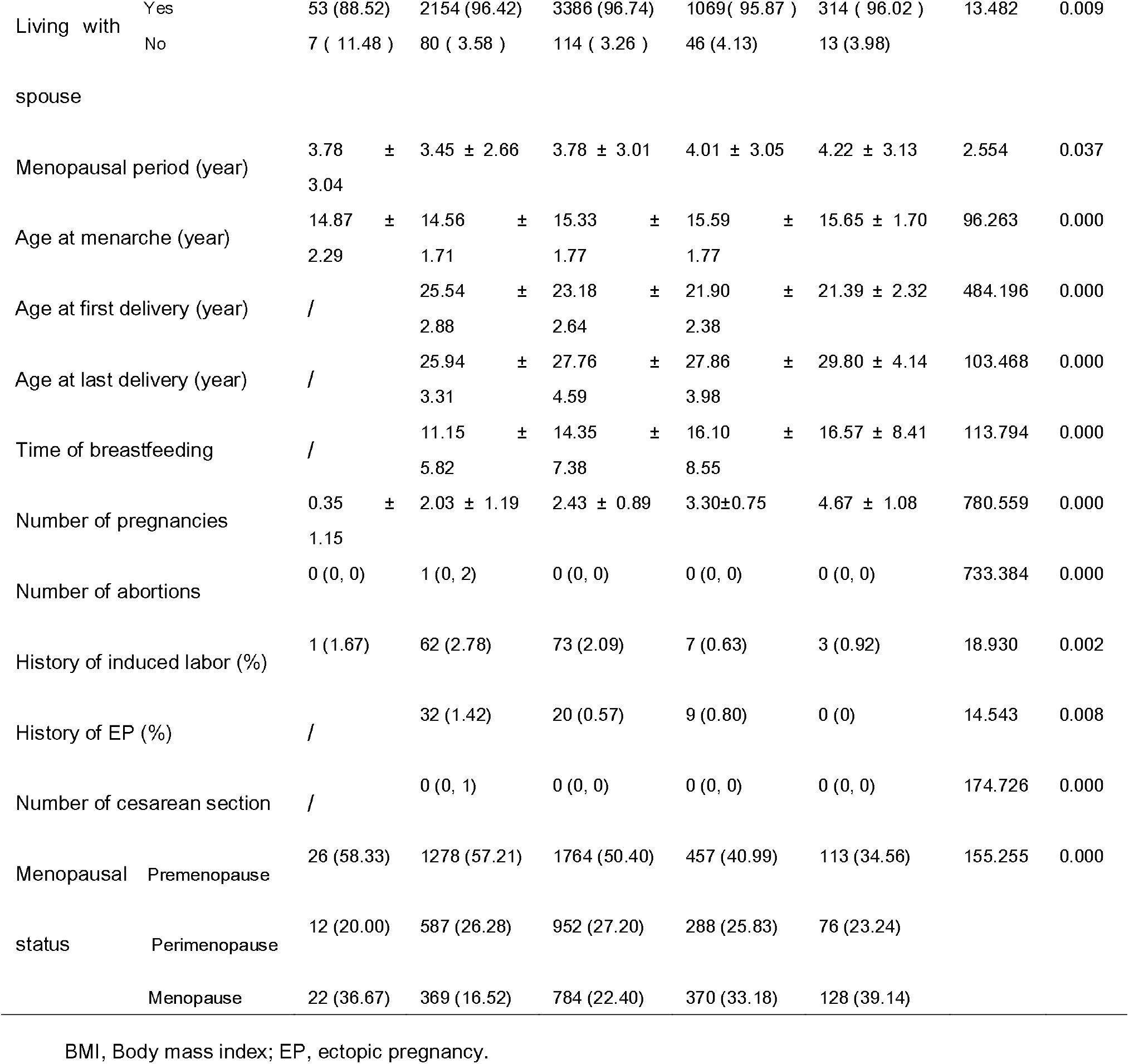
Baseline characteristics of study participants with different numbers of births

### Effects of the number of births on the age at menopause

The Kaplan–Meier analysis indicated that the median age at menopause was 48 years and the mean age at menopause was 47.91 ± 3.31 years. Moreover, 1673 women experienced menopause. A trend of increased mean age at menopause was observed with the increased number of births (*F* = 4.794; *P* = 0.001). The median age at menopause was 3 years earlier among women with nulliparity than among women with ≥4 births (median: 46 vs 49). Women with 1 birth had a higher risk of menopause (HR = 1.322; 95% CI: 1.072–1.629). After adjustment for confounding factors, no relationship was observed between the number of births and the age at menopause (*P* = 0.595) (Table 2).

**Table 2.**
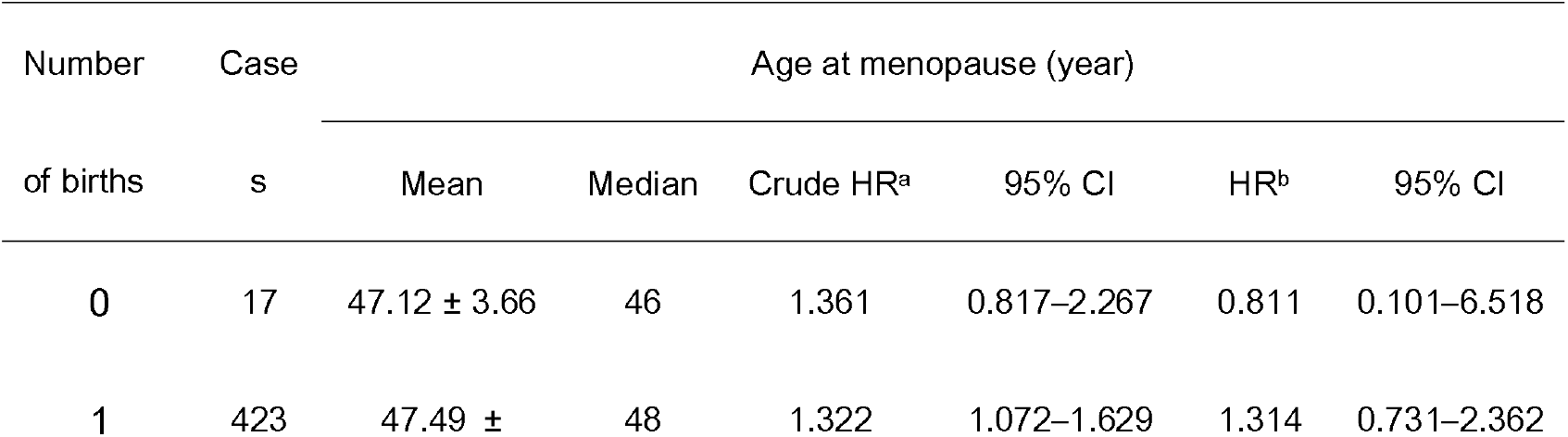
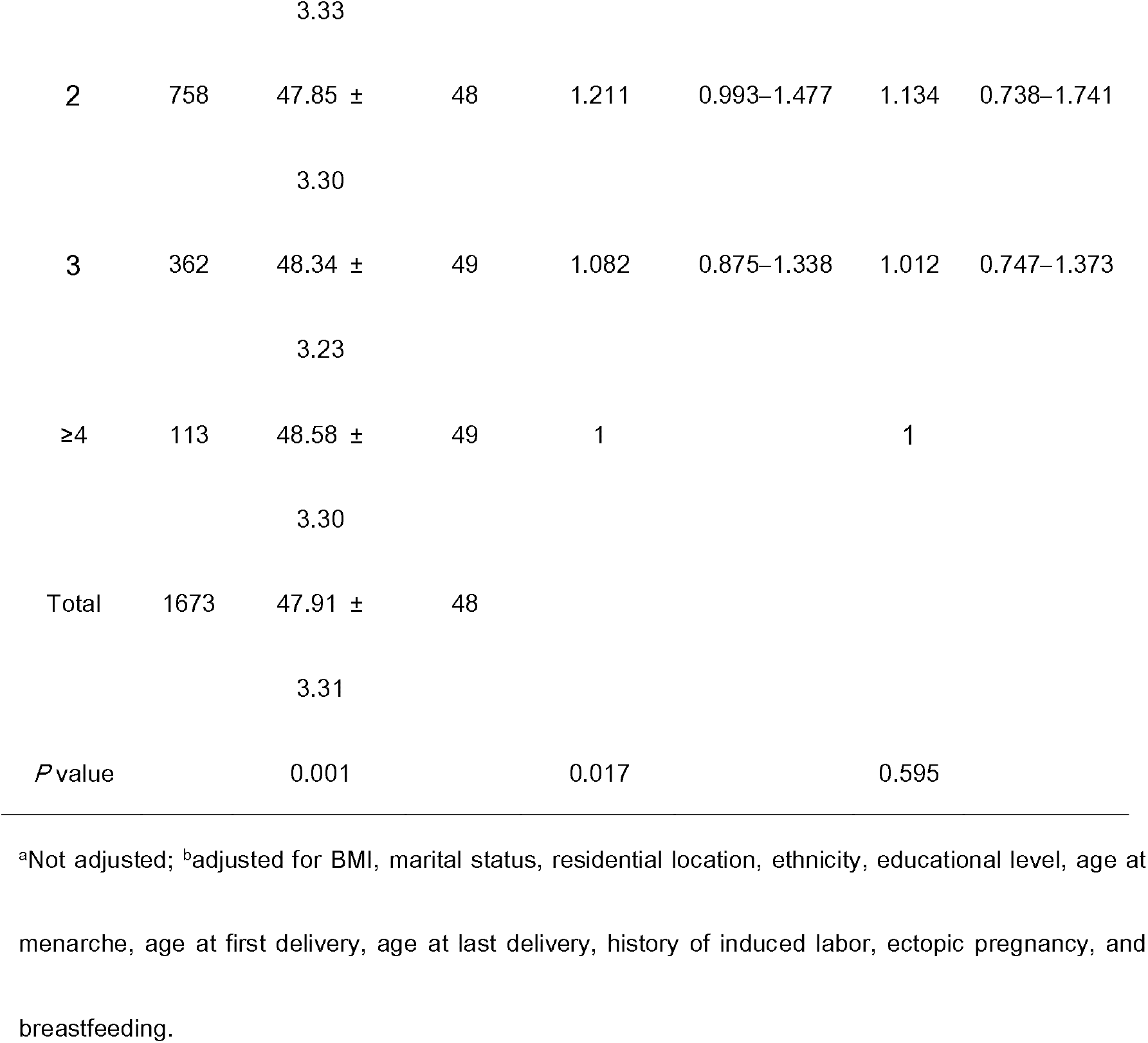
Effects of the number of births on the age at menopause

The Kaplan–Meier estimates are shown in Figure 1. Women with higher number of births had a higher risk of menopause (*χ*^2^ = 12.042; *P* =0.017). However, the Cox regression analysis showed that the relationships were not significant after adjustment for body mass index (BMI), marital status, residential location, ethnicity, educational level, age at menarche, age at menopause, age at first delivery, age at last delivery, history of induced labor, ectopic pregnancy, and breastfeeding (*P* = 0.488)(Figure 2).

**Figure 1.**
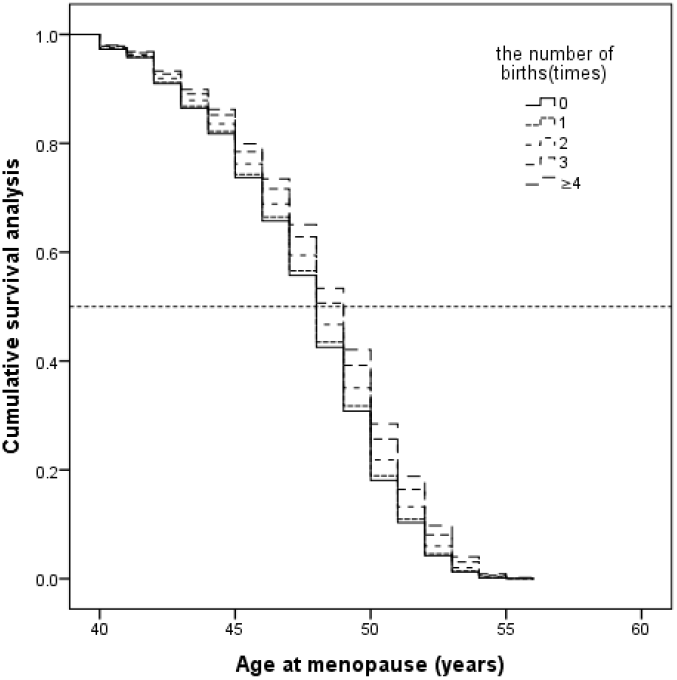
Kaplan–Meier curve of the effect of parity on the age at menopause. Women with higher number of births had a higher risk of menopause (*χ*^2^ =12.042; *P* = 0.017)

**Figure 2.**
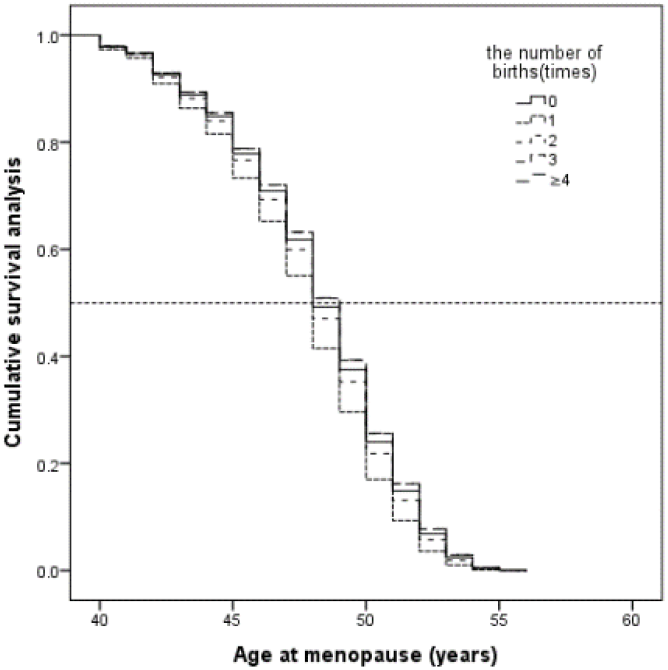
Kaplan–Meier curve of the effect of births on the age at menopause adjusted for BMI, marital status, residential location, ethnicity, educational level, age at menarche, age at first delivery, age at last delivery, history of induced labor, ectopic pregnancy, and breastfeeding, as revealed by the Cox regression analysis (*P* = 0.488)

### Relationship between parity and the severity of menopausal syndrome

Figure 3 shows that, excluding nulliparous women, the rest of the group had a significant increase in the moderate and severe menopausal syndrome with a higher number of births (*χ*^2^ = 289.079; P = 0.000). The proportions of moderate (50.5%) and severe syndrome (17.1%) were highest in women with 4 births. The prevalence of severe syndrome was higher in nulliparous women than in those with 1 or 2 births (8.3% vs. 4.3% and 7.6%).

**Figure 3.**
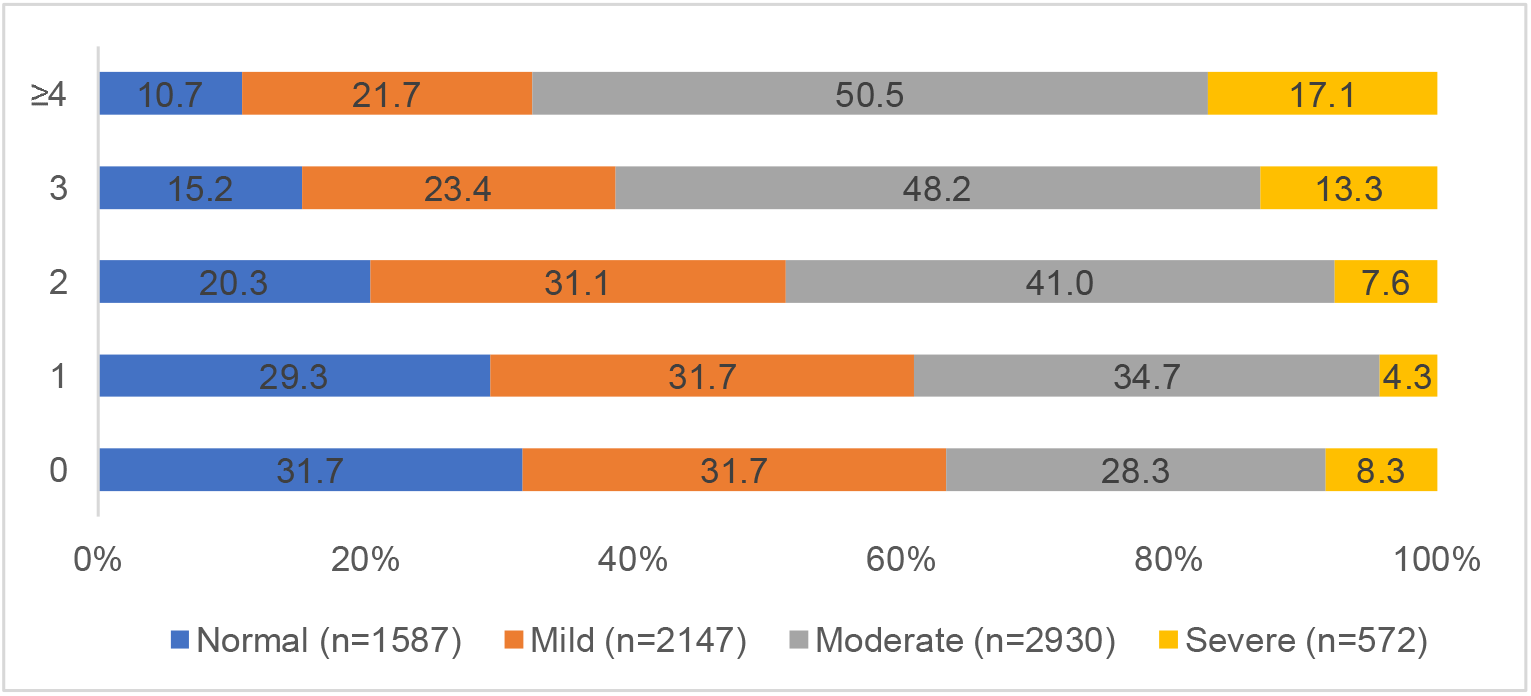
Relationship between the incidence of menopausal syndrome and the number of births.

Table 3 shows the findings of univariable logistic regression analysis. After adjustment for confounding factors, a lower risk of moderate and severe syndrome was found among women with 1 birth (OR = 0.534; 95% CI: 0.330–0.863) and those with 1 birth (OR = 0.360; 95% CI: 0.187–0.693) and 2 births (OR = 0.540; 95%CI: 0.317–0.922), respectively, compared with those with ≥4 births.

**Table 3.**
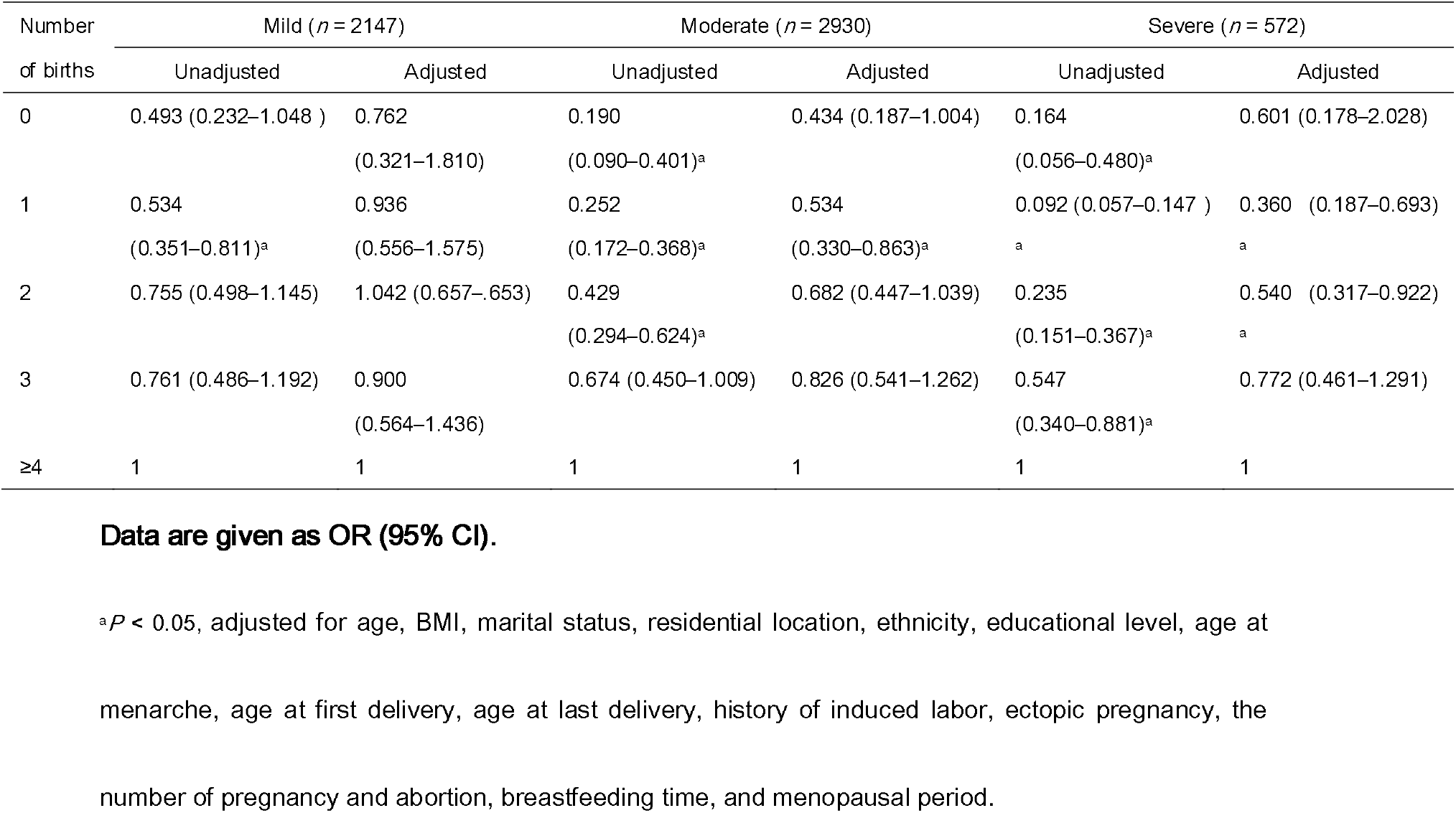
Multivariable Logistic regression analysis of the effects of the number of births on the menopausal syndrome

### Relationship between parity and three main menopausal syndromes

The risk of somatic and psychological symptoms increased with the higher number of births, but the difference was not found to be statistically significant on applying the multivariable logistic regression analysis. Compared with ≥4 births, women with 1 and 2 births had a lower risk for urogenital symptoms (Table 4).

**Table 4.**
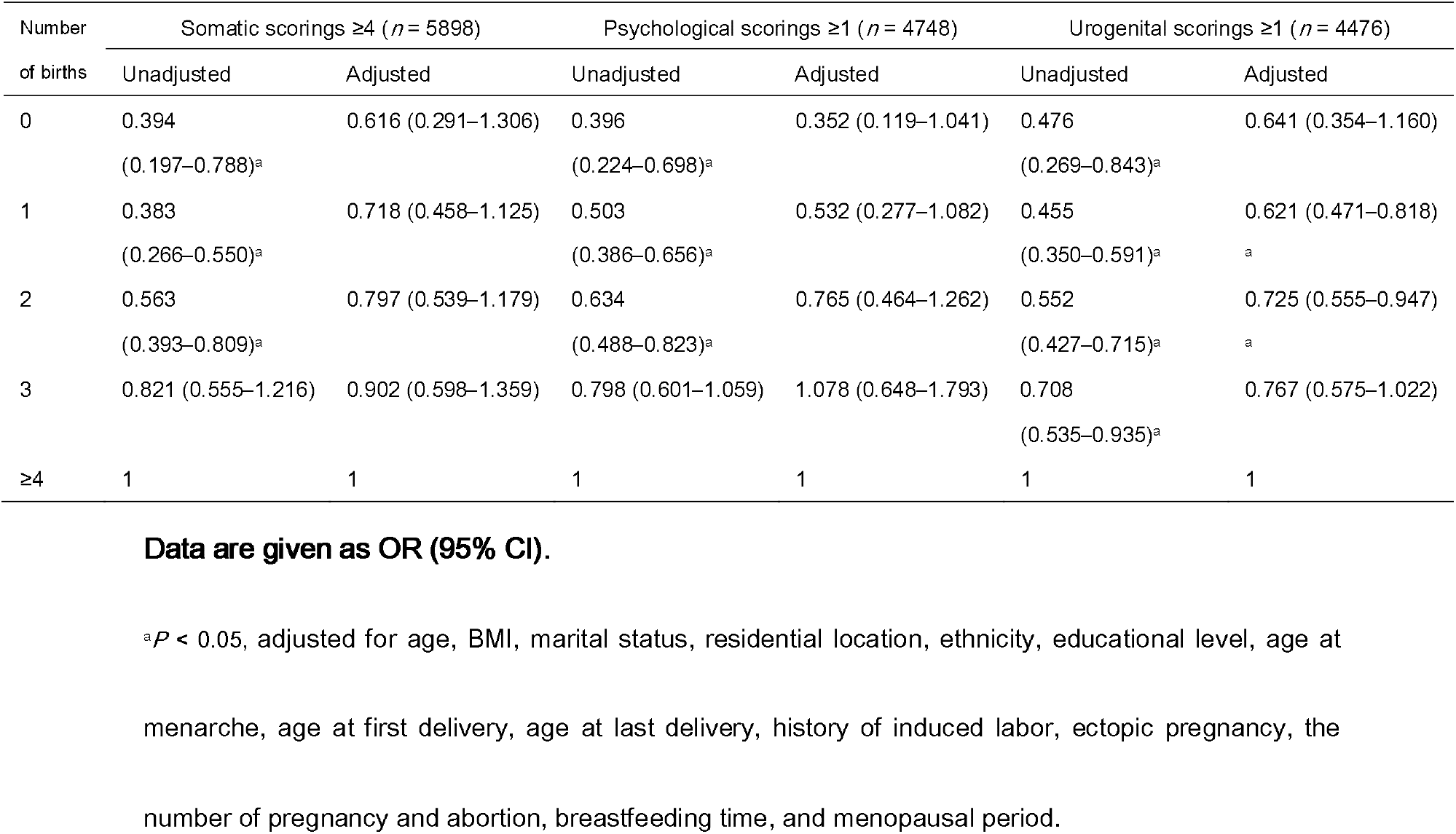
Relationship between parity and three main menopausal syndromes, as revealed by univariable and multivariable logistic regression analyses

## Discussion

This study evaluated the relationship between delivery times and the age at menopause and menopausal syndrome among Chinese women in Gansu. Although the mean age at menopause was associated with the increased number of births, the relationships between birth times and age at menopause were not found to be significant on applying the Cox regression analysis. Women with nulliparity and more births seemed to have higher risks of moderate and severe menopausal syndrome. However, the potential beneficial effects of 1 or 2 births on menopausal syndromes were observed by applying multivariable logistic regression analysis, particularly in urogenital symptoms.

### Relationship between parity and the age at menopause

Menopause is an important milestone for midlife women because it represents the loss of fertility and the transition to postreproductive life. In the present study, the mean age at menopause was 47.91 years, which was a little earlier compared with that in other provinces in China[4] and developed countries, such as Europe and Australia [16] [17] [18; 19], America[20], and Korea[21]. It was speculated that the participant (40–55 years) bias might have partly led to the results. Meanwhile, Gansu Province is located in Northwest China, where the economic level is worse than that in the eastern and southern regions in China. Numerous factors, including genetic, environmental, and lifestyle-related, influence the onset of menopause.

Compared with other studies, this study showed that the relationship between parity and age at menopause was attenuated by adjustment for confounding factors, which was consistent with the findings of Rizvanovic and colleagues [6]. However, other studies found that higher parity was associated with older age at menopause[22] and nulliparity was associated with increased risk of premature and early menopause[4; 23]. One of these studies showed that nulliparous women were at more than fivefold increased risk of premature menopause and twofold increased risk of early menopause compared with women who had two or more children[23]. It was speculated that parity and fewer cumulative menstrual cycles in parous women might be associated with a larger reserve of oocytes and longer exposure to estrogen[24]. Importantly, it was found that infertile patients had similar AMH levels and AFC compared with controls with no history of infertility in an age-adjusted linear regression analysis[25], confirming the results of the present study. However, the study participants were aged less than 40 years. It was believed that ethnicity, living environment, economy, geography, diet characteristics, smoking, physical activity, individual characteristics, and sample size might be the factors leading to inconsistent results. This study was based on the province’s large sample size, representing the overall situation in Gansu Province, and the results were credible.

### Relationship between parity and severity of menopausal symptoms

The present study showed that the risk of moderate and severe menopausal syndrome increased with ≥3 births; nulliparous women had a higher risk of severe syndrome compared with those having 1 or 2 births. Indeed, the results of the study were in agreement with previous findings indicating that multiparity aggravated menopausal symptoms [4; 8; 26]. Several potential reasons accounted for this relation, one of which was that parous women were prone to living in rural areas and having low income and low education level. Therefore, it was quite possible that they could not get enough postpartum nursing and adequate nutrition after giving birth. Meanwhile, multiparous women were at a younger age at first gravidity and delivery. Therefore, the organ systems were not able to meet the increased physiological demands of pregnancy in younger mothers[27]. It was also harder to raise more children due to the lack of financial support and mothers needing more labor. Moreover, infertile women were significantly more likely to report severely decreased libido and more than twice as likely to report severe vaginal dryness[23], which was similar to the findings of the present study.

In addition, the present study found that the risk of somatic and psychological symptoms was not statistically significantly different after adjustment for several confounding factors. A lower risk was found in women with one and two births compared with nulliparous and parous (3 and ≥4 births) women. Previous studies on the factors associated with reproduction and menopausal symptoms indicated that the number of deliveries was truly related to urogenital symptoms. Botlero and colleagues reported that the risk of developing stress incontinence increased with obesity and parity[28]. Women scoring positive on the infertility index were significantly more likely to report severely decreased libido and more than twice as likely to report severe vaginal dryness in multivariable models[29]. Yeniel and colleagues showed that vaginal delivery was an independent risk factor for prolapse with an OR of 2.92 (95% CI: 1.19–7.17) compared with nulliparity. Also, each vaginal delivery increased the risk of prolapse (OR = 1.23; 95% CI: 1.12–1.35) after controlling for all confounding factors[30]. In addition, multiparity had an obvious impact on urinary incontinence[31], vaginal atrophy, and lower urinary tract symptoms[32]. Awwad and colleagues found that clinically significant pelvic organ prolapse (POP) was found in 3.6% of nulliparous, 6.5% of primiparous, 22.7% of secondiparous, 32.9% of triparous, and 46.8% of tetraparous women[33]. It was thought that the determinants of experiencing menopausal symptoms were complex, representing biological, psychological, and social factors. The prevalence of sexual problems and urinary symptoms in the present study was 48.89% and 33.04%, respectively, both accounting for 61.86% (not shown in the tables), which was lower than the prevalence of 79.1%–83% reported in other studies [34; 35]. The European REVIVE survey, which included the largest cohort of postmenopausal women studied to date, confirmed that vulvovaginal atrophy was frequent but still underdiagnosed and undertreated[36].

A large number of factors result in an independent relationship between parity and urogenital symptoms. A previous study demonstrated that vaginal delivery was at a greater risk of presenting with UI because of structural changes in the pelvic floor and repeated injury to the muscles, nerves, and connective tissue of the pelvic floor during childbirths[37; 38]. Further, the participants in the present study had a low education level and resided in economically backward areas. Therefore, they lacked postpartum care and guidance, leading to sexual problems and urinary symptoms [39]. Therefore, improving the medical level and accelerating development are essential to achieving better health and quality of life in women.

Investigations on birth limits have been conducted in China for 40 years. The nulliparity women are usually infertile or choose to remain childless. Meanwhile, women with multiple births reside in remote rural areas, have a lower educational level, and live with physical labor. Therefore, the higher number of births is possibly a result of low income and educational background, which decide the higher rate of menopausal syndrome and are not the reason for the severe menopausal syndrome. This may due to the fact that women with higher parity are often found in low-income families with low educational background [40]. These observations were also consistent with the results of the present study. Meanwhile, adverse outcomes of more children worsen the economic burden and need more efforts to earn a living. This is why the findings of this study result were not similar to previous findings.

This study, being a cross-sectional study, had several potential limitations, including recollection bias. Information about the number of births and the age at menopause was obtained using self-reported questionnaires, leading to recall bias. The study did not exclude all the confounding effects of the natural aging process that might influence the experience of symptoms. Also, a validity check for the self-reported questionnaire could not be conducted. Further randomized, controlled, prospective studies are needed to substantiate the relationship between multiparity and menopausal syndrome.

## Conclusions

This study suggested a relationship between parity and the age at menopause and menopausal syndrome, which was particularly specific to Chinese women in Gansu. Together, the data both supported and extended the current understanding of the individual number of births as a factor contributing to a woman’s experience of menopausal symptomology. However, further higher-quality prospective studies should be conducted to confirm this relationship between parity and menopause, which might serve as a valid tool for practitioners to consider when counseling their premenopausal patients about the incidence and severity of menopause.

## Data Availability

The data in this article comes from the survey. All the data are reliable. Please contact the author to get the data.

## Contributors

Xuehong Zhang and Rui Zhang were responsible for the study concept and design.

Xiaoyan Sun and Weiguo Li were responsible for statistical analysis of the results and wrote the manuscript.

Xiping Shen, Yongbin Lu and Yiqing Wang contributed to data analysis.

Rui Zhang, Liyan Wang, Xiaoyan Sun, Junxia An, Xiaorong Luo and Haiying Zhu conducted the survey.

## Ethical approval

Informed consent was obtained from all individual participants in the study. All authors approved the final manuscript. The Medical Ethical Committee of the First Hospital of Lanzhou University approved the study protocol.

## Research data (data sharing and collaboration)

There are no linked research data sets for this paper. Data will be made available upon request.

## Abbreviations

KMI: the Kupperman Menopause Index
HR: hazard ratio
CI: confidence interval
OR: the odds ratio.

## References

[1] Ebtekar, F., Dalvand, S., and Gheshlagh, R. G. The prevalence of metabolic syndrome in postmenopausal women: A systematic review and meta-analysis in Iran. Diabetes & metabolic syndrome 2018;

[2] Karvonen-Gutierrez, C. A., Park, S. K., and Kim, C. Diabetes and Menopause. Current diabetes reports 2016; 16:20

[3] Newson, L. Menopause and cardiovascular disease. Post reproductive health 2018; 24:44–49

[4] Li, L., Wu, J., Pu, D., Zhao, Y., Wan, C., Sun, L., Shen, C. E., Sun, W., Yuan, Z., Shen, Q., et al. Factors associated with the age of natural menopause and menopausal symptoms in Chinese women. Maturitas 2012; 73:354–360

[5] Dorjgochoo, T., Kallianpur, A., Gao, Y. T., Cai, H., Yang, G., Li, H., Zheng, W., and Shu, X. O. Dietary and lifestyle predictors of age at natural menopause and reproductive span in the Shanghai Women’s Health Study. Menopause 2008; 15:924–933

[6] Rizvanovic, M., Balic, D., Begic, Z., Babovic, A., Bogadanovic, G., and Kameric, L. Parity and menarche as risk factors of time of menopause occurrence. Medical archives (Sarajevo, Bosnia and Herzegovina) 2013; 67:336–338

[7] Klingberg, S., Brekke, H. K., Winkvist, A., Engstrom, G., Hedblad, B., and Drake, I. Parity, weight change, and maternal risk of cardiovascular events. American journal of obstetrics and gynecology 2017; 216:172.e171–172.e115

[8] Choi, J. I., Han, K. D., Kim, S. J., Kim, M. J., Shin, J. E., and Lee, H. N. Relationship between delivery history and health-related quality of life in menopausal South Korean women: The Korea National Health and Nutrition Examination Surveys. Maturitas 2016; 92:24–29

[9] Nanri, A., Mizoue, T., Noda, M., Goto, A., Sawada, N., and Tsugane, S. Menstrual and reproductive factors and type 2 diabetes risk: The Japan Public Health Center-based Prospective Study. Journal of diabetes investigation 2018;

[10] Guan, H. B., Wu, Q. J., and Gong, T. T. Parity and kidney cancer risk: evidence from epidemiologic studies. Cancer epidemiology, biomarkers & prevention : a publication of the American Association for Cancer Research, cosponsored by the American Society of Preventive Oncology 2013; 22:2345–2353

[11] Guo, P., Xu, C., Zhou, Q., Zhou, J., Zhao, J., Si, Z., Shen, C., and Feng, C. Number of parity and the risk of gallbladder cancer: a systematic review and dose-response meta-analysis of observational studies. Archives of gynecology and obstetrics 2016; 293:1087–1096

[12] Harlow, S. D., Gass, M., Hall, J. E., Lobo, R., Maki, P., Rebar, R. W., Sherman, S., Sluss, P. M., and de Villiers, T. J. Executive summary of the Stages of Reproductive Aging Workshop +10: addressing the unfinished agenda of staging reproductive aging. Climacteric : the journal of the International Menopause Society 2012; 15:105–114

[13] Freeman, E. W., and Sherif, K. Prevalence of hot flushes and night sweats around the world: a systematic review. Climacteric : the journal of the International Menopause Society 2007; 10:197–214

[14] Islam, M. R., Gartoulla, P., Bell, R. J., Fradkin, P., and Davis, S. R. Prevalence of menopausal symptoms in Asian midlife women: a systematic review. Climacteric : the journal of the International Menopause Society 2015; 18:157–176

[15] Monteleone, P., Mascagni, G., Giannini, A., Genazzani, A. R., and Simoncini, T. Symptoms of menopause - global prevalence, physiology and implications. Nature reviews Endocrinology 2018; 14:199–215

[16] Schoenaker, D. A., Jackson, C. A., Rowlands, J. V., and Mishra, G. D. Socioeconomic position, lifestyle factors and age at natural menopause: a systematic review and meta-analyses of studies across six continents. International journal of epidemiology 2014; 43:1542–1562

[17] Dratva, J., Gomez Real, F., Schindler, C., Ackermann-Liebrich, U., Gerbase, M. W., Probst-Hensch, N. M., Svanes, C., Omenaas, E. R., Neukirch, F., Wjst, M., et al. Is age at menopause increasing across Europe? Results on age at menopause and determinants from two population-based studies. Menopause 2009; 16:385–394

[18] Bjelland, E. K., Hofvind, S., Byberg, L., and Eskild, A. The relation of age at menarche with age at natural menopause: a population study of 336 788 women in Norway. Hum Reprod 2018; 33:1149–1157

[19] Stepaniak, U., Szafraniec, K., Kubinova, R., Malyutina, S., Peasey, A., Pikhart, H., Pajak, A., and Bobak, M. Age at natural menopause in three central and eastern European urban populations: the HAPIEE study. Maturitas 2013; 75:87–93

[20] Gold, E. B., Crawford, S. L., Avis, N. E., Crandall, C. J., Matthews, K. A., Waetjen, L. E., Lee, J. S., Thurston, R., Vuga, M., and Harlow, S. D. Factors related to age at natural menopause: longitudinal analyses from SWAN. American journal of epidemiology 2013; 178:70–83

[21] Lim, H. S., Kim, T. H., Lee, H. H., Park, Y. H., Kim, J. M., and Lee, B. R. Hypertension and age at onset of natural menopause in Korean postmenopausal women: Results from the Korea National Health and Nutrition Examination Survey (2008-2013). Maturitas 2016; 90:17–23

[22] Gold, E. B. The timing of the age at which natural menopause occurs. Obstetrics and gynecology clinics of North America 2011; 38:425–440

[23] Mishra, G. D., Pandeya, N., Dobson, A. J., Chung, H. F., Anderson, D., Kuh, D., Sandin, S., Giles, G. G., Bruinsma, F., Hayashi, K., et al. Early menarche, nulliparity and the risk for premature and early natural menopause. Hum Reprod 2017; 32:679–686

[24] Santoro, N., Brockwell, S., Johnston, J., Crawford, S. L., Gold, E. B., Harlow, S. D., Matthews, K. A., and Sutton-Tyrrell, K. Helping midlife women predict the onset of the final menses: SWAN, the Study of Women’s Health Across the Nation. Menopause 2007; 14:415–424

[25] Hvidman, H. W., Bentzen, J. G., Thuesen, L. L., Lauritsen, M. P., Forman, J. L., Loft, A., Pinborg, A., and Nyboe Andersen, A. Infertile women below the age of 40 have similar anti-Mullerian hormone levels and antral follicle count compared with women of the same age with no history of infertility. Hum Reprod 2016; 31:1034–1045

[26] Ceylan, B., and Ozerdogan, N. Menopausal symptoms and quality of life in Turkish women in the climacteric period. Climacteric : the journal of the International Menopause Society 2014; 17:705–712

[27] Gibbs, C. M., Wendt, A., Peters, S., and Hogue, C. J. The impact of early age at first childbirth on maternal and infant health. Paediatric and perinatal epidemiology 2012; 26 Suppl 1:259–284

[28] Botlero, R., Davis, S. R., Urquhart, D. M., Shortreed, S., and Bell, R. J. Age-specific prevalence of, and factors associated with, different types of urinary incontinence in community-dwelling Australian women assessed with a validated questionnaire. Maturitas 2009; 62:134–139

[29] Nelson, D. B., Sammel, M. D., Patterson, F., Lin, H., Gracia, C. R., and Freeman, E. W. Effects of reproductive history on symptoms of menopause: a brief report. Menopause 2011; 18:1143–1148

[30] Yeniel, A. O., Ergenoglu, A. M., Askar, N., Itil, I. M., and Meseri, R. How do delivery mode and parity affect pelvic organ prolapse? Acta obstetricia et gynecologica Scandinavica 2013; 92:847–851

[31] Juliato, C. R., Baccaro, L. F., Pedro, A. O., Gabiatti, J. R., Lui-Filho, J. F., and Costa-Paiva, L. Factors associated with urinary incontinence in middle-aged women: a population-based household survey. International urogynecology journal 2017; 28:423–429

[32] Maserejian, N. N., Curto, T., Hall, S. A., Wittert, G., and McKinlay, J. B. Reproductive history and progression of lower urinary tract symptoms in women: results from a population-based cohort study. Urology 2014; 83:788–794

[33] Awwad, J., Sayegh, R., Yeretzian, J., and Deeb, M. E. Prevalence, risk factors, and predictors of pelvic organ prolapse: a community-based study. Menopause 2012; 19:1235–1241

[34] Palma, F., Volpe, A., Villa, P., and Cagnacci, A. Vaginal atrophy of women in postmenopause. Results from a multicentric observational study: The AGATA study. Maturitas 2016; 83:40–44

[35] Nappi, R. E., and Kokot-Kierepa, M. Vaginal Health: Insights, Views & Attitudes (VIVA) - results from an international survey. Climacteric : the journal of the International Menopause Society 2012; 15:36–44

[36] Nappi, R. E., Palacios, S., Panay, N., Particco, M., and Krychman, M. L. Vulvar and vaginal atrophy in four European countries: evidence from the European REVIVE Survey. Climacteric : the journal of the International Menopause Society 2016; 19:188–197

[37] Thom, D. H., and Rortveit, G. Prevalence of postpartum urinary incontinence: a systematic review. Acta obstetricia et gynecologica Scandinavica 2010; 89:1511–1522

[38] Handa, V. L., Harris, T. A., and Ostergard, D. R. Protecting the pelvic floor: obstetric management to prevent incontinence and pelvic organ prolapse. Obstetrics and gynecology 1996; 88:470–478

[39] Coyne, K. S., Wein, A., Nicholson, S., Kvasz, M., Chen, C. I., and Milsom, I. Economic burden of urgency urinary incontinence in the United States: a systematic review. Journal of managed care pharmacy : JMCP 2014; 20:130–140

[40] Nisar, N., Sikandar, R., and Sohoo, N. A. Menopausal symptoms: prevalence, severity and correlation with sociodemographic and reproductive characteristics. A cross sectional community based survey from rural Sindh Pakistan. JPMA The Journal of the Pakistan Medical Association 2015; 65:409–413

